# Savor the flavor — a randomized double-blind study on taste-enhanced placebo analgesia in healthy volunteers

**DOI:** 10.1101/2021.08.16.21262058

**Authors:** Matthias Zunhammer, Gerrit Goltz, Maximilian Schweifel, Boris A. Stuck, Ulrike Bingel

**Affiliations:** Department of Neurology and Center for Translational Neuro- and Behavioral Sciences (C-TNBS), University Hospital Essen, Hufelandstr. 55, 45147 Essen, Germany; Department of Otolaryngology, Head and Neck Surgery, University Hospital Marburg, Philipps-Universität Marburg, Marburg, Germany

**Keywords:** placebo effect, placebo analgesia, taste, gustatory, human, experimental pain, cold pain, cold pressor test, cold pressor task, heart rate, thermal pain, treatment context, contextual treatment effect

## Abstract

We conducted a randomized, double-blind, between-group study to investigate how the taste of oral medication affects placebo analgesia. Over three sub-studies, 318 healthy volunteers (297 included) were subjected to experimental tonic cold water pain (cold pressor test) before and after receiving taste-neutral (water), bitter (quinine), sweet (saccharine), or no placebo drops. Pain ratings indicated that taste enhances placebo analgesia. This effect was small but accounted for a substantial portion of the overall placebo effect and was comparable to WHO stage 1 analgesic effects. Moreover, placebo treatments were associated with an increase in peak heart rate response to cold water. Adverse effects were minimal. These results indicate that added taste may be an easy-to-implement, cost-effective, and safe way to optimize treatment outcomes and that taste-neutral preparations may reduce placebo-related outcome variance in clinical trials. Further studies are needed to test if these findings can be translated into clinical scenarios.

## Background

Placebo effects are increasingly recognized as powerful modulators of health and treatment outcomes ^1^. Placebo effects are not limited to inert placebo treatments but also contribute substantially to active treatments ^2^; this effect is particularly large for pain and depression where up to 70% of overall treatment outcomes may be attributed to placebo effects ^1^. These discoveries call for a systematic exploitation of placebo effects in clinical care to enhance the efficacy of gold-standard treatments ^3^. Extensive research over the past decades has linked placebo effects to expectancy, learning and social cognition mechanisms ^4^, which are driven by various aspects of the treatment context, ranging from the information delivered along with a treatment ^5^, to features of the treatment itself, such as labeling ^6^, color ^7^, and even price ^8^.

Taste is a prominent characteristic of any oral medication and has been suggested to modulate placebo effects^9,10^. Studies in humans and rodents have indicated that pharmacologically induced immunosuppression can be re-evoked by presenting a conditioned taste, equivalent to a ‘learned placebo effect’ ^11^. Yet, although placebo analgesia is the best studied form of the placebo effect ^12^, surprisingly little is known about the impact of gustatory sensations. This lack of knowledge is unfortunate, considering the dissatisfactory situation in many chronic pain settings, where established analgesics often show limited efficacy (e.g. see NSAIDS against chronic lower back pain ^13^), since adding flavor to oral medication could be an cost-effective and easy-to-implement way to utilize placebo effects in clinical care. Moreover, a better understanding of the gustatory component of placebo effects may be useful to improve the efficacy of clinical trials by minimizing placebo-related variability ^14,15^.

Here we performed a series of three randomized, double-blind studies in a total of 318 healthy volunteers to assess whether bitter and sweet flavor (compared to neutral flavor and no treatment) enhance placebo effects on experimental pain. The experimental design is outlined in Fig. 1. Pain was induced using the cold pressor task (CPT), an established experimental model of tonic pain^16,17^. Pre-versus post-treatment pain ratings served as the primary outcome. Pain tolerance, adverse effects of the (placebo) treatment and HR-responses were assessed as secondary outcomes. Further, the influence of participants’ treatment expectation, subjective taste intensity and/or taste valence ratings were assessed.

**Fig. 1:**
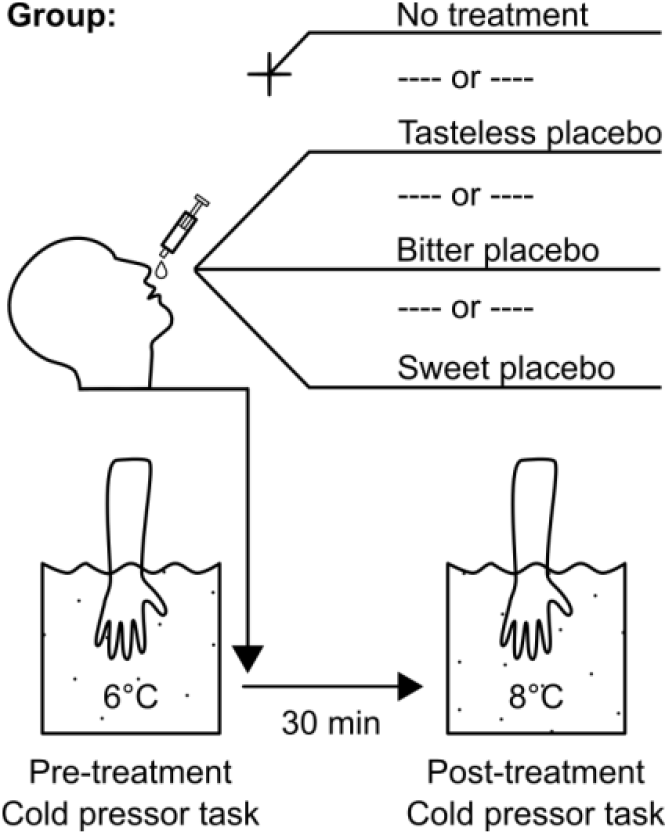
Experimental design. Across 3 sub-studies, participants were allocated to either receive no treatment, or tasteless placebo, or bitter placebo, or sweet placebo, after baseline testing. Water-temperature was covertly increased by 2 °C before post-treatment testing to simulate a weak analgesic treatment effect, as is typical for clinical settings.

## Results

Healthy participants of both sexes were allocated to either receive no treatment, tasteless placebo, bitter placebo, or sweet placebo. We recruited, allocated and tested 318 participants across the three sub-studies; twenty-one participants (6%) were excluded from the main analysis based on pre-defined exclusion criteria, yielding an effective sample of 297 (Table 1, Tables S1, Supplementary Methods). The established cold pressor test (CPT)^18^ was used to induce experimental pain. CPT was performed before and 30 minutes after placebo treatment, or before and after a corresponding waiting period in the no treatment group. In all groups, a weak analgesic background treatment effect was simulated^19^ by covertly increasing CPT water temperatures from 6.0 °C at baseline to 8.0 °C at post-treatment to simulate a actual medication effect.

**Table 1:** Sample descriptives, pooled across sub-studies.

| **Group** | **No treatment** | **Taste-neutral placebo** | **Bitter placebo** | **Sweet placebo** | **Total:** |
| --- | --- | --- | --- | --- | --- |
| **Randomized (n)** | 81 | 81 | 81 | 75 | 318 |
| **Excluded (n)<sup>a</sup>** | 10 | 6 | 2 | 3 | 21 |
| **Per-protocol sample (n)<sup>b</sup>** | 71 | 75 | 79 | 72 | 297 |
| **Heart Rate Recordings (n)<sup>c</sup>** | 68 | 71 | 74 | 67 | 280 |
| **Sex (%male)** | 49% | 45% | 51% | 60% | 51% |
| **Age, mean (SD)** | 24.3 (2.7) | 24.4 (3.4) | 24.6 (3.4) | 24.4 (3.2) | 24.5 (3.2) |
| **Handedness (%right)** | 92% | 91% | 97% | 96% | 94% |

<sup>a</sup> Reasons for exclusion were: previous participation in similar studies (n = 5), alcohol consumption on the day before testing (n = 4), lack of pain response at CPT-baseline (n = 3), dizziness/hypotension in response to CPT-testing (n = 2), psychiatric diagnose (n = 2), surgery within the last 6 months, or use of analgesic medication within the last week (n = 3), an endocrine condition (n = 1), or technical failure during testing (n = 1).
<sup>b</sup> An additional intention-to-treat analysis was performed with all available data.
<sup>c</sup> Number of participants where both pre- and post-treatment heart rate recordings could be analyzed. Reasons for additional exclusion compared to the per-protocol sample were: recording failure (n = 16) and extreme values (n = 1).

### Primary outcome: Pain ratings in the cold pressor test

In the CPT, participants submerged their hand into cold water for as long as tolerable, or a safety maximum of 180 seconds, while continuously rating pain intensity on a visual analog scale (VAS), using a mechanical sliding lever. Based on the rating-curves obtained during the CPT (Fig. S1) the primary outcome area-under-the-pain-curve (AUPC) was calculated according to Koltzenburg et al. (2006)^16^. A higher AUPC indicates higher individual pain sensitivity: an AUPC of 0% denotes a constant VAS pain rating of 0, whereas an AUPC of 100 % denotes the immediate termination of testing due to pain intolerance, or equivalently, a constant VAS rating of 100 for 180 seconds.

Mean pain rating curves and changes in AUPC from pre-to post-treatment are shown in Fig. 2 (also see: Table S2). A general linear model (GLM) analysis was performed to estimate effects of factors *group* on post-treatment AUPC. Baseline pre-treatment values were included in the model as a covariate^20^ to account for inter-individual differences in pain sensitivity. The fixed factor *study* was included to account for potential differences between sub-studies (see: Tables 2). The factor of interest *group* explained ∼6% of residual variance in post-treatment AUPC, which is considered a small-to-moderate effect in statistical terms^21^ (Table 2a). As expected, the covariate *pre-treatment AUPC* was the best predictor of post-treatment values, explaining most of the variance (86%) in post-treatment AUPC (Table 2a), justifying its use as a baseline control. The factor sub-study explained little variance (1%) indicating that mean sub-study differences played a minor role in treatment related changes. GLM-contrasts indicated that placebo treatment (pooled: neutral, bitter, sweet) was superior to no treatment, reducing AUPC by an estimated -5.31%, 95%CI [-8.19, -2.78] (β = -0.20, 95% CI [-0.30, -0.11], t(292) = -3.87, p < .0001); 6.9 participants needed to be treated with *any* placebo to achieve an additional responder (defined as -30% pain reduction from baseline) over no treatment. This is considered a small effect in both clinical^22^ and statistical^21^ terms. Individually, all three placebos groups showed a small benefit (standardized effect size: ∼0.2 standard deviations^21^) over to the no-treatment group (see: Table 2b).

**Fig. 2:**
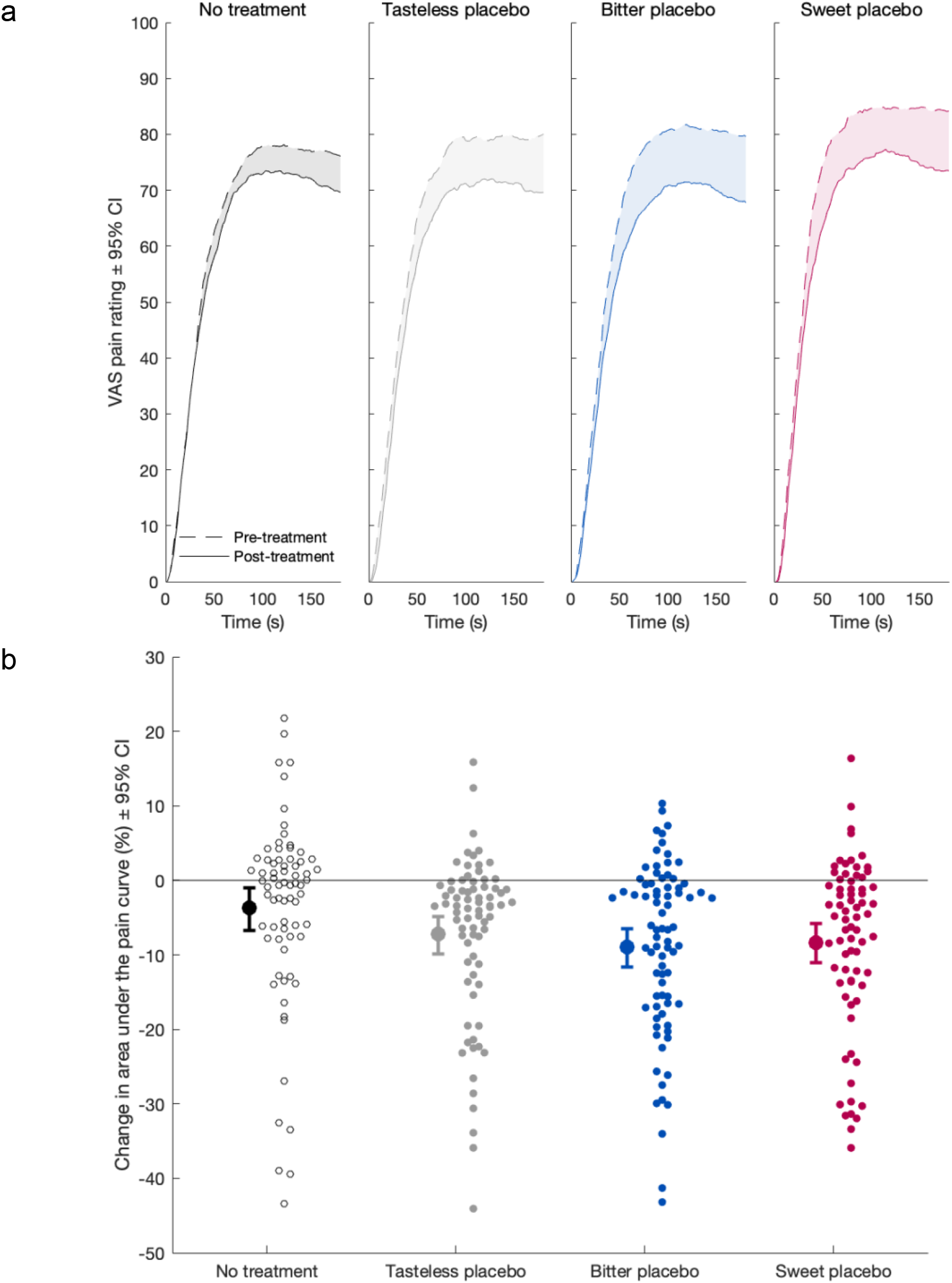
analgesic effects of placebo treatment on a) continuous pain ratings and b) area under the pain rating curve (AUPC) a) Mean pain rating curves obtained during Cold Pressor Test (CPT) for pre-treatment at 6°C (dashed lines) and post-treatment at 8°C (solid lines) timepoints. Ratings of participants terminating testing before the maximum of 180 s, were carried forward with a pain rating of 100. Differences in pain rating curves over time are highlighted as an area and correspond to the means in Fig. 2b. b) Means ± bootstrapped 95 % confidence intervals (BCa) of area under the pain rating curve in %, shown next to individual data points (n = 297). Negative values indicate that post-treatment pain ratings were lower than pre-treatment ratings.

**Table 2a:**
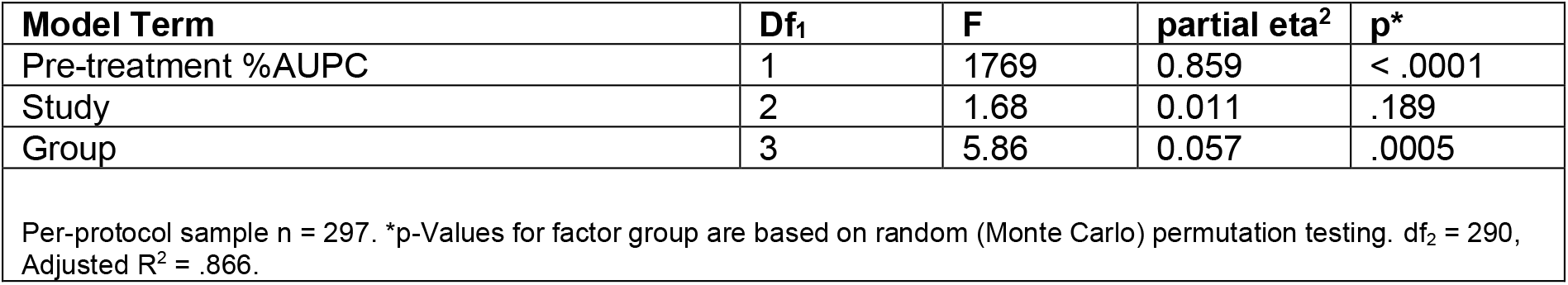
GLM results of % area under the pain curve.

**Table 2b:** GLM contrasts of factor *group*, for % area under the pain curve.

| Model Term | B [95% CI]* | Beta [95% CI]* | t | p** |
| --- | --- | --- | --- | --- |
| Neutral Placebo > No Treatment | -3.55 [-7.06, -0.75] | -0.13 [-0.26, -0.02] | -2.16 | .069 |
| Bitter Placebo > No Treatment | -5.77 [-9.39, -2.77] | -0.21 [-0.35, -0.11] | -3.54 | .003 |
| Sweet Placebo > No Treatment | -6.76 [-10.9, -3.62] | -0.25 [-0.40, -0.13] | -3.53 | .0004 |
| Bitter Placebo > Neutral Placebo | -2.22 [-5.34, 0.96] | -0.08 [-0.20, 0.04] | -1.38 | .246 |
| Sweet Placebo > Neutral Placebo | -3.21 [-6.83, 0.03] | -0.12 [-0.25, 0.00] | -1.68 | .095 |
| Sweet Placebo > Bitter Placebo | -0.99 [-4.80, 2.32] | -0.04 [-0.18, 0.08] | -0.52 | .612 |

Per-protocol sample n = 297. \* 95% Confidence Intervals based on bootstrapping (asymmetric, BCa method). \*\*p-Values are based on random (Monte Carlo) permutation testing. B denotes unstandardized model coefficients (unit: %AUPC), beta values show standardized model coefficients (unit: SD). df<sub>2</sub> = 290, Adjusted R<sup>2</sup> = .866.

Flavored placebo groups (pooled) showed an additional AUPC reduction by -2.57%, 95% CI [-5.35, 0.26] (β = -0.09, 95% CI[-0.19, 0.01], t(291) = -1.80, p = .125) over the tasteless placebo group; 7.2 participants needed to be treated with flavored placebo to achieve an additional responder. This is considered a very small absolute effect in both clinical^22^ and statistical^21^ terms. Nevertheless, the taste-enhancement effect amounted to +72% (95% CI [+151%, -7.3%]) of the observed placebo effect in the taste-neutral group, or equivalently, a boost-factor of 1.72. Also independently, bitter and sweet placebo treatment groups showed an additional benefit versus neutral placebo (standardized effect size: ∼0.1 standard deviations, Table 2b), with sweet placebo showing a marginal advantage over bitter placebo (Table 2b).

Auxiliary analyses were performed to corroborate these findings (also see: Supplementary Results, Fig. S2): Of note, effect size estimates and statistical test results were confirmed, when repeating the analysis with all participants tested, including those fulfilling the pre-defined exclusion criteria (Table S3), which largely excludes that deliberate selection bias affected analysis. Moreover, directions of effect and effect sizes were essentially confirmed, when separately analyzing maximum pain tolerance time in the subgroup of participants terminating testing early (Tables S4) and average pain rating in the subgroup that did not terminate testing (Tables S5), instead of using AUPC as a summary measure.

### Secondary outcome II: adverse effects

Adverse effects attributed to the (placebo) treatment were assessed as another secondary outcome. Levels of adverse effects were very low across the sample (Table S8). Most participants (n=191, 64% of the per-protocol sample) did not report any placebo-related side effects and no single rating exceeded “moderate” severity. Two participants reported dizziness and/or showed signs of hypotension in response to CPT-testing after completing the experimental session. Most frequent side effect was “drowsiness” (n = 65), followed by “feeling hot” (n = 28) and “palpitations” (n = 24). Average side-effect scores were very low (0.8 ± 1.2 units out of 78 units possible) and there were no detectable differences in side effects scores between placebo groups (F[2, 218] = 0.15, p = .859, partial eta^2^ = 0.0007), even when only considering participants that showed any side effects (F[2, 89] = 0.08, p = .925, partial eta^2^ = 0.0009).

### Secondary outcome: heart rate

CPT typically induces a spike in heart rate (HR)^23,24^. These increases in HR are a compensatory cardiac response^25^ to the cold-stressor, and have been shown to reflect autonomic nervous system responsivity, i.e. sympathetic activation and parasympathetic de-activation^24,26,27^. Here, we assessed treatment group effects on the CPT HR-response as a secondary outcome measure that may also shed light on the mechanisms and physiological effects of placebo treatment. To this end, we obtained continuous pulse spectrophotometer-based HR recordings during CPT and assessed the peak CPT HR-response (observed HR maximum during CPT, minus mean HR during a 15-seconds pre-CPT baseline). Valid HR-recordings were available for 280 out of the 297 participants in the per-protocol sample (Table 1); 17 participants could not be analyzed due to missing or invalid recordings. Single-participant HR-curves during CPT are provided in Figure S3, descriptive results are provided in Table S6.

Mean continuous HR-curves during CPT and CPT HR-peak amplitudes are shown in Fig. 3. The main GLM tested effects of factor *group* on post-treatment peak CPT HR-response, controlling for pre-treatment peak CPT HR-response, and the fixed factor *study*. ANCOVA indicated that treatment group explained ∼7% of residual variance in post-treatment AUPC, which is considered a small-to-moderate effect size. (Table 3a). In the no-treatment group, peak HR responses to the 2nd CPT were clearly reduced compared to the 1^st^ CPT (Fig. 3), which is expected as the 2^nd^ CPT was a weaker stressor (water bath was +2 °C warmer) and since HR-responses to CPT are known to attenuated with repeated exposure ^23^. Contrarily, placebo treatments increased peak CPT HR-response over no treatment by +2.92 bpm (95%CI [0.91, 4.91], β = 0.34, 95% CI [0.10, 0.56], t(275) = 2.78, p = .006). In particular, flavored placebo groups showed increased CPT HR-responses compared to neutral placebo (+3.23 bpm, 95%CI [1.09, 5.17], β = 0.38, 95% CI [0.16, 0.57], t(274) = 2.94, p = .005), with a pronounced effect in the bitter placebo group (Table 3b). These results indicate that flavored placebo treatments, particularly bitter treatment, increase HR-responses to the cold-water challenge relative to neutral tasting placebos. Several auxiliary analyses were performed to corroborate these finding: In short, no pre- or post-treatment group differences in baseline HR were detected and results were replicated when the repeating the analysis with all participants tested, and (s. Supplementary Results, Table S7). Of note, there was no appreciable relationship between pre-treatment peak CPT HR-response and %AUPC (see: Figure S4) suggesting that peak CPT HR-response are not a surrogate marker of pain, replicating previous findings ^28,29^.

**Fig. 3:**
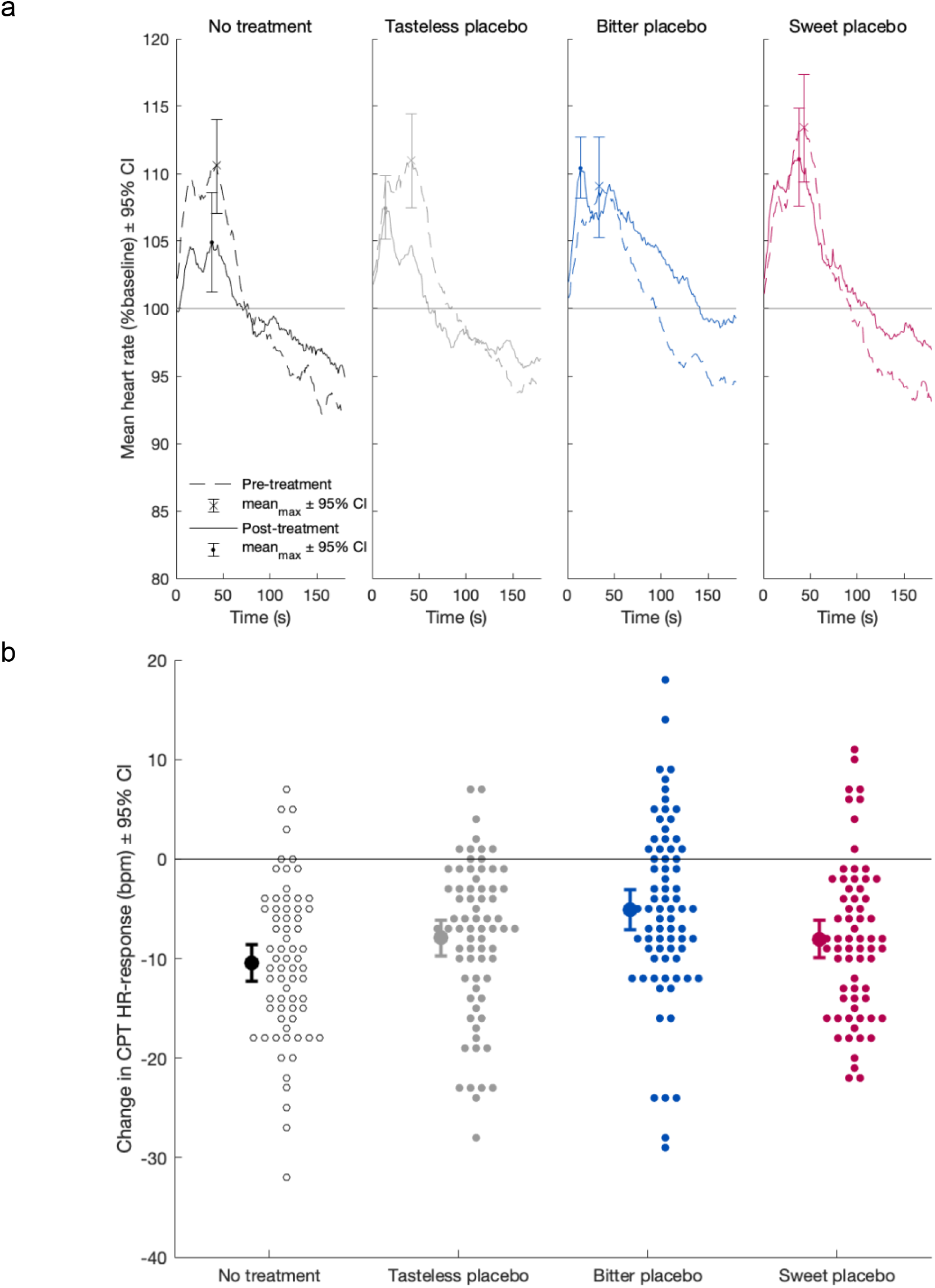
effects of placebo treatment on a) continuous heart rate recordings during the cold pressor test (CPT) and b) change in CPT heart rate response (pre vs post-treatment) a) Mean heart-rate curves obtained during Cold Pressor Test (CPT) for pre-treatment at 6°C (dashed lines) and post-treatment at 8°C (solid lines) timepoints. Maxima of mean HR curve are highlighted ± bootstrapped 95 % confidence intervals (BCa). b) shown next to individual data points (n = 280). Negative values indicate that cold-pressor-induced heart rate peaks were lower post-treatment compared to pre-treatment. b) Means ± bootstrapped 95 % confidence intervals (BCa) of change in CPT heart-rate response from pre-to post-treatment timepoints, shown next to individual data points (n = 280). Negative values indicate that the post-treatment heart rate response (maximum HR-peak during CPT) was smaller than pre-treatment response.

**Table 3a:** GLM results of CPT HR-response.

| Model Term | Df<sub>1</sub> | F | partial eta<sup>2</sup> | p* |
| --- | --- | --- | --- | --- |
| Pre-treatment %AUPC | 1 | 58.0 | 0.175 | < .0001 |
| Study | 2 | 1.31 | 0.010 | .270 |
| Group | 3 | 6.41 | 0.066 | .00018 |

Per-protocol sample (w valid HR recordings) n = 280. \*p-Values for factor group are based on random (Monte Carlo) permutation testing. df<sub>2</sub> = 273, Adjusted R<sup>2</sup> = .205.

**Table 3b:** GLM contrasts of factor *group*, for CPT HR-response.

| Model Term | B [95% CI]* | Beta [95% CI]* | t | p** |
| --- | --- | --- | --- | --- |
| Neutral Placebo > No Treatment | 0.92 [-1.43, 3.37] | 0.11 [-0.17, 0.4] | 0.74 | .069 |
| Bitter Placebo > No Treatment | 4.99 [2.57, 7.53] | 0.58 [0.3, 0.89] | 4.04 | .0004 |
| Sweet Placebo > No Treatment | 2.7 [-0.18, 5.31] | 0.32 [-0.03, 0.62] | 1.88 | .246 |
| Bitter Placebo > Neutral Placebo | 4.07 [1.73, 6.54] | 0.47 [0.2, 0.75] | 3.34 | .095 |
| Sweet Placebo > Neutral Placebo | 1.78 [-1.16, 4.55] | 0.21 [-0.13, 0.54] | 1.24 | .612 |
| Sweet Placebo > Bitter Placebo | -2.29 [-5.67, 0.79] | -0.27 [-0.65, 0.09] | -1.58 | .612 |

Per-protocol sample (w valid HR recordings) n = 280. \* 95% Confidence Intervals based on bootstrapping (asymmetric, BCa method). \*\*p-Values are based on random (Monte Carlo) permutation testing. B denotes unstandardized model coefficients (unit: beats-per-minute), beta values show standardized model coefficients (units: SD). df<sub>2</sub> = 273, Adjusted R<sup>2</sup> = .205.

### Intervention checks and auxiliary analyses

To aid the interpretation of potential placebo and taste effects, we obtained and explored participant ratings of treatment expectations, subjective taste intensity, and taste valence. Expectations of pain relief were obtained after treatment, but before the post-treatment CPT, using a visual analogue scale (VAS) asking for “What extent of pain relief do you expect from treatment” with endpoints labeled,, 0: no relief” and,, 100: very strong pain relief”. Expectations of analgesia were moderate on average (41.9 β 20.2, Table S8) and the factor group did not explain sizeable amounts of variance (F[2, 221] = .304, p = .738, partial eta^2^ = 0.0014, corrected for fixed sub-study effects).

Retrospective taste ratings were obtained after post-treatment CPT, using a VAS asking “How strong was the flavor of the medication:”, with endpoints labeled,, 0: no taste” and,, 100: very intense taste. Taste intensity ratings strongly differed between levels of factor *group* (F[2, 221] = 65.6, p < .001, eta^2^ = 0.238), with higher taste intensity ratings in the bitter (b = +30.4 95% CI [24.5, 36.3], β = +1.21 95% CI [0.98, 1.45], t = 10.1, p < .001), and sweet (b = +9.1 95% CI [1.7, 16.5], β = +0.36 95% CI [0.07, 0.66], t = 2.41, p = .017), compared to the tasteless placebo group (7.6 ± 10.3, Table S8). Of note, taste intensity ratings in the bitter group were elevated compared to the sweet group (b = +21.2 95% CI [13.8, 28.7], β = +0.85 95% CI [0.55, 1.15], t = 5.58, p < .001), indicating that sweet and bitter conditions were not fully equivalent in terms of recalled taste intensity, despite two pilot studies that aimed at balancing taste intensity (Figure S5).

Retrospective taste valence ratings were obtained on a VAS (“How pleasant/unpleasant was the flavor of the medication:” ranging from: -50: “very unpleasant”, via 0: “neutral taste”, to +50: “very pleasant taste”) also strongly differed between groups (F[2, 221] = 33.7, p < .001, eta^2^ = 0.152). On average, the taste of placebo medication was recalled as neutral in the tasteless group (mean = -3.6 ± 9.2, Tables S8), as moderately unpleasant in the bitter group (b = -9.91 95% CI [-14.3, -5.50], β = -0.52 95% CI [-0.74, -0.29], t = -4.40, p < .001), and as moderately pleasant in the sweet group (b = +13.3 95% CI [7.70, 18.8], β = +0.69 95% CI [0.40, 0.98], t = 4.67, p < .001). Taken together, these results indicate that our three placebo interventions successfully induced beliefs of pain relief and successfully evoked different gustatory experiences. An exploratory analysis of potential associations of %AUPC with ratings of treatment expectations, taste intensity, or taste valence merely indicated weak relationships (Tables S9).

## Discussion

In this large-scale, experimental proof-of concept study in healthy volunteers, we assessed the effects of flavoured vs unflavoured placebo treatments against pain. Several important findings derive from our study: (i) Bitter and sweet placebo drops were 1.8 and 2.5 times more effective in reducing pain than neutral placebo, (ii) placebo treatment caused minimal adverse effects, regardless of taste, and (iii) placebo treatments, especially the bitter placebo, enhanced the cardiac response to cold pressor stress.

Our results show that the analgesic efficacy of oral placebo medication can be enhanced by adding flavor and, more generally that sensory experiences that accompany medical treatments can enhance placebo effects. The estimated additional benefit of taste-enhanced placebo treatment (as compared to neutral placebo) was limited when compared to the effect sizes typically reported in CPT experiments for opioids ^16,30–33^, ketamine ^32^, and pregabalin ^32^, yet comparable to the effect sizes reported for several non-steroidal anti-inflammatory analgesics ^30,32–34^. Importantly, adding flavor to the placebo treatment enhanced the response rate (more than -30% pain reduction from baseline), with 7.2. participants needed to obtain an additional responder. Although this is considered a small effect, it should be noted that an NNT of 7 is comparable to first line treatments such as pregabalin or gabapentin used for neuropathic pain, which highlights its potential for clinical use at comparable efficacy^35^. Contrary to tried-and-tested drugs, taste-enhanced placebo drops may provide additional analgesic effects with minimal risk, side-effects, or costs. Considering the dissatisfactory situation in many chronic pain settings, where established analgesics often show limited efficacy^35^ (e.g. see NSAIDS against chronic low back pain ^13^), our present results highlight a potential ‘low hanging fruit’ for additional patient benefit. Further trials are needed to test whether our findings can be translated to verum analgesics, and whether clinical populations can benefit from flavoredflavoured gold-standard verum analgesics. Moreover, taste enhancement may be of relevance for open-label placebo treatments, which are increasingly recognized to have clinically relevant effects in pain disorders ^36–38^, depression ^39^ or cancer related fatigue ^40^. Based on previous findings ^41^ we are optimistic that the reported effect may be even larger in patients.

The overall placebo effect on pain ratings observed in this study was small, while previous experimental placebo studies typically report effect sizes of d = 1.0 (standard deviations) ^42^. This difference can be explained by the fact that most experimental placebo studies rely on within-subject designs ^42^ and include conditioning procedures to enhance the magnitude and sustainability of placebo analgesia ^43^. Here, we deliberately chose a between-subject design and induced placebo analgesia through verbal information only. This decision was based on several reasons: Firstly, within-subject comparisons of treatment conditions may affect gustatory perception and introduce biases in judgment and decision-making, or ‘demand characteristics’ as participants are able to directly compare treatments ^44^. Secondly, we wanted to use a placebo setting that is ready-to-implement into clinical routine. While the experience of treatment efficacy, as induced in conditioning protocols, can boost treatment responses^19^, the translation of such strategy into clinical settings can be difficult, given that often no efficient treatment is available to induce a positive treatment experience. Thirdly, we wanted to keep our study comparable to clinical trial settings, where participants are typically naïve to a novel treatment. These design choices distinguishes our study from earlier studies in the field of placebo research that e.g. achieved immunomodulation via taste stimuli^11,45^.

Besides its use as a model of experimental pain, the cold pressor test is an established stress-test for cardiovascular- and autonomic nervous system function^46^. After CPT onset, biomarkers of sympathetic NS activity typically increase and biomarkers of parasympathetic NS activity decrease in the first minute after CPT onset, returning to, or below baseline thereafter ^24,26,27,46–49^. Here, we assessed the amplitude of peak HR-increases after CPT onset vs pre-CPT baseline^50^. We not only replicated the known CPT-induced HR-peak, but also found that the peak HR-response to CPT differed between treatment groups. The no treatment group showed diminished HR-responses in the second CPT, while placebo treatment groups, especially the bitter treatment group, showed a sustained, high HR-response in both the first and second (post-treatment) CPT session (Fig. 3). These results are remarkable for two reasons. First, to date only few studies could demonstrate placebo effects on physiological, cardiac outcome parameters in a sizeable sample ^12,51^.

Second, the observed placebo-induced changes in cardiac reactivity are interesting regarding the physiological mechanisms associated with placebo analgesia. Flavored placebo treatment groups showed both increased CPT-induced HR-responses and increased placebo analgesia. However, no direct relationship between individual HR-responses and individual placebo analgesia was found, which indicates that the two effects may be independent. Thus, our findings link placebo treatment, but not necessarily placebo analgesia to increased cardiac autonomic nervous system responses under cold pressor stress. The detailed mechanisms and causality of this observed placebo effect on HR-responses are unclear to date. It is interesting to speculate that the observed autonomic cardiac response may reflect a general defensive and/or self-regulatory response to placebo treatment. In the context of a clinical trial setup, a novel medical oral treatment, may pose as a mild stressor keeping participants autonomous nervous system alert. Of note, brain stem regions, such as the PAG, have been associated with responses to threatening stimuli^52^ and with placebo analgesia^53^ (including descending pain modulation), and coordinate autonomic responses^54^.

A number of additional findings from our study could provide further insights into the underlying mechanisms of these taste-enhanced placebo effects: Differences between sweet and bitter placebo conditions were too small to be estimated with sufficient confidence. Moreover, the participants’ recall of taste valence indicated that the subjective pleasantness or unpleasantness of the treatment showed no sizeable relationship with pain ratings. These results suggest that taste-related placebo effects in adults are not primarily driven by the hedonic qualities of the treatment, which may distinguish tasted-enhanced placebo effects from the analgesic effects of sweet taste observed in neonates ^55^. We would nevertheless argue for using pleasant, rather than unpleasant flavors in future studies with active drugs, as the taste of oral medication may impact treatment outcomes beyond placebo effects. An aversive taste may simply reduce the willingness to adhere to the treatment regimen or even induce nausea or other adverse effects and thus directly affect the efficacy of verum analgesics ^56,57^.

Although sizeable expectations of pain relief were reported by our participants in all placebo conditions, placebo taste had no additional effect on conscious expectations of pain relief. Further, individual treatment expectations showed no association with placebo analgesia. These results indicate that the taste-related differences in placebo efficacy between groups are insufficiently explained by conscious treatment-related beliefs and support the notion that placebo effects may be induced by sub-conscious mechanisms ^58,59^.

These findings have to be seen in the light of several limitations. The present study cannot be considered confirmatory according to the null-hypothesis significance testing paradigm, as the study sample size was extended in two steps (Sub-Studies 2 and 3), after Sub-Study 1 turned out to be inconclusive on its own. Future replication experiments are therefore necessary to confirm these findings. Data were obtained in a controlled experimental setting with inert placebo treatments and with an established cold-water pain induction method used in healthy volunteers. It is unclear whether our results translate to verum treatments and/or clinical pain populations, especially since altered nociceptive, as well as gustatory processes may pertain in chronic pain disorders. Moreover, the taste perception does not simply depend on the choice of gustatory qualities, such as sweet, bitter, sour, salty, savory, on taste intensity and on taste valence (unpleasant/pleasant), but also depends on other factors, most prominently retronasal olfaction ^60^. Our present study was limited to two moderately intense taste qualities, namely bitter and sweet in comparison to tasteless placebo drops. Future experiments with a wider range of taste preparations are desirable to better understand the importance of other taste dimensions for the placebo effect.

## Conclusion

Our present study results indicate the taste of a medication is a contextual treatment factor that affects the analgesic efficacy of placebo treatment in healthy volunteers ^61^. Given that placebo effects are not limited to placebo medication, but also affect active pharmacological treatments ^62^, our findings may have implications for clinical routine and clinical trials. If our results translate to clinical settings, adding taste to oral analgesic medication may be a save and inexpensive way to improve clinical outcomes. Although the observed effect sizes indicate that the added benefit of flavor may be modest, the cost-effectiveness and minimal risk involved may justify its use. By contrast, avoiding detectable flavor in both verum and placebo medication may be instrumental to minimize placebo-induced outcome variance in clinical trials and treatment-related stress in research volunteers. Future trials have to confirm whether these findings can be translated to verum analgesics and whether clinical populations can benefit from flavour-based enhancements of oral analgesics.

## Methods

### Ethics and participants

The present study was conducted in accordance with the Declaration of Helsinki, approved by the ethics committee of the Universitätsklinikum Essen (16-7163-BO and amendments). The study was registered in the German Clinical Trials Register (DRKS00011688), after the start of data collection. Healthy, young (age: 18-40 years) participants were recruited at the University of Duisburg-Essen by advertisement on bulletin boards. Participants were informed in written and verbal form and signed consent was obtained in every case. Pre-defined exclusion criteria were: history of neurological, psychiatric, or major internal disorders, Raynaud syndrome, injuries to the upper limbs, history of recurrent cramps or syncopes, pregnancy, acute infections, alcohol consumption within the last 48 hour, and use of analgesic or psychotropic substances within the last two weeks. All candidate participants were tested, regardless of exclusion criteria, as long as testing was deemed safe. The decision on study exclusion/inclusion was made after testing, before analysis, by MZ and UB; these measures were taken to reduce sampling bias and allow for estimating sampling-bias in an intention-to-treat analysis. Participants received a compensation of 50 € for their participation.

### Study design - Randomization

The present study encompasses three subsequent sub-studies. Each study followed the same experimental protocol, and each study followed a randomized, double-blind parallel group design. For each sub-study a separate randomization list was created a-priori by author MZ, who used Microsoft Excel’s RAND function to generate a random, sequential list of participant numbers with group allotments. Group allotment to participant numbers was balanced against participant sex in sub-studies 1 and 2, and random in sub-study 3. Participants were sequentially assigned participant numbers according to their first scheduled experimental visit by the experimenters (authors GG and MS). MZ had no influence on (and no a-priori knowledge of) assignments of individual participant numbers to participants. The experimenters and the study participants in the placebo treated groups were blind in respect to group allocation. For the no-treatment groups, no blinding could be achieved, as the knowledge of *not* being treated was essential for this experimental condition.

### Study design - Groups

Sub-Study 1 was planned as a confirmatory study, with a target sample size of n = 138 (n = 128, plus up to 10 dropouts) and an allocation ratio of 1:1:1 to groups “no-treatment”, “taste-neutral placebo”, and “bitter placebo”. The sample size for Sub-Study 1 was pre-determined in a power analysis that was built on the assumption of a large placebo effect (Cohen’s d = 1.0 according to a meta-analysis of experimental placebo studies^42^) and consequently assumed small effect sizes of modulatory taste. After analysis of Sub-Study 1, this assumption turned out to be untenable as the observed main placebo effect was far from large (d = 0.54). We consequently decided to extend the sample post-hoc by adding additional sub-studies. The sample size for these subsequent studies was not determined by *a-priori* statistical power calculations but determined according to the available resources for testing. Sub-study 2 was identical to Sub-Study 1 in terms of study-design, aimed at recruiting 30 (50% male) additional participants, and was conducted by author MS. In Sub-Study 3, a ‘sweet placebo’ condition was introduced to allow for a wider generalizability of results (in terms of taste) and to allow for further mechanistic insights (i.e. to explore whether placebo effects are driven by the hedonic qualities of taste ^9,10^). Sub-Study 3 aimed at recruiting 150 (50% male) eligible participants. Participants were randomly assigned to one of four groups that either received no treatment, tasteless placebo, bitter placebo, or sweet placebo, with an allocation ratio of 1:1:1:3, respectively. Testing was conducted by author MS.

### Placebo treatment

Participants were informed that the purpose of the study was to investigate “interaction effects between a tried-and-tested analgesic drops and the individual genetic background”. Participants were not informed that the focus of the study was taste perception.^1^ We chose oral drops as the mode of administration, because drops entail reliable gustatory stimulation and because drops are a common route of administration of several potent analgesics (e.g. Tilidin, Tramadol, Metamizol). A bitter taste was chosen in Sub-Studies 1 and 2 since oral medications are commonly associated with a bitter taste ^56,57^.

Placebo solutions were prepared by a certified pharmacist and stored at 4 °C. For the tasteless treatment group, the drops consisted of purified water. An aqueous quinine solution (0.8 mM/l, 0.03g Quinine-dihydrochloride in 100 ml purified water) was used in the bitter placebo group, as commonly used in gustatory research ^63^. The exact dose was chosen according to pilot experiments, with the aim to elicit an intensely bitter taste that disappears within 30 minutes (Fig. S5). The total dose of 0.24 mg quinine given to participants in the bitter taste group, corresponds to the amount of quinine contained in 3 to 4 ml of a typical off-the-shelf,, *Tonic Water*” ^64^ and is unlikely to have noteworthy physiological effects. For the sweet treatment group, an aqueous saccharine solution (1.0 mM/l, 0.02g Na-Saccharin in 100 ml purified water) was used ^65^. The dose was chosen according to pilot experiments with the aim to elicit an intensely sweet taste that approximately matches the quinine solution in terms of taste intensity at intake and disappears within 30 minutes (Fig. S5). The total dose of 0.16 mg saccharin given to participants in the sweet group, corresponds to the amount of saccharin contained in 2 to 3 ml of a typical off-the-shelf,, *Soft Drink* “^66^ and is unlikely to have noteworthy physiological effects.

For each participant number, MZ prepared sequentially numbered 1.0 ml plastic syringes with 0.8 ml placebo solution on the day of application, the type of placebo solution was determined according to the randomization list. Aside from the numbering, placebo containers were not discernible. Drops were stored at 4 °C until administration by the experimenters. Participants were instructed by the experimenters to retain the drops sublingually for 10 seconds, to disperse the drops within the mouth subsequently, and to swallow the remainder after another 10 seconds. In all treatment groups, the experimenter applied the 0.8 ml of placebo drops, sublingually.

### Testing schedule

Data collection took place from Dec 19^th^, 2016 to Mar 07^th^ 2017 (Study 1, first participant allocated Dec 16^th^, 2016), from Mar 26^th^ 2018 to Apr 13^th^ 2018 (Sub-Study 2), and from Apr 25^th^ 2018 to Jul 05^th^ 2018 (Sub-Study 3). The testing schedule was identical for all three Sub-Studies: After participants arrived at our laboratories at Essen University Hospital, informed consent was obtained, the current health status was examined, and potential exclusion criteria were recorded. Participants were introduced to the CPT and visual analog scale (VAS) rating procedures according to a standardized protocol. A first CPT was performed as a pre-treatment baseline. Then, oral placebo drops were applied in the treatment groups, while the non-treatment received no treatment. Subsequently, a 30 min waiting period was observed in all groups, during which participants were asked to fill in questionnaires. The waiting period was implemented a) to simulate the delay-of-onset of typical analgesic drugs, b) to avoid confounding subsequent testing procedures with ongoing gustatory stimulation (e.g. through distraction), and c) to allow the skin to recover from pre-testing CPT. After the waiting period, expectations of treatment-induced pain relief were obtained in the treatment groups using a VAS (,,0: no relief“,,, 100: very strong pain relief“) shown on-screen. Subsequently, the post-treatment CPT was performed. After the CPT, participants in the treatment groups were asked to provide VAS ratings regarding the medication’s a) overall efficacy (,,0: no effect”, 100: very strong pain relief), b) taste intensity (,,0: no taste”,,, 100: very intense taste“), and c) taste pleasantness (,,-50: very unpleasant taste“,,, 0: taste-neutral“,,, 50: very pleasant taste“). A blood sample was taken after testing to allow for genetic assessments in future studies. Lastly, participants were asked for treatment side effects and discharged after debriefing. Treatment-related side effects were queried systematically using a questionnaire that covers the 17 common side effects of over-the-counter analgesics and placebo medication ^67^. For each side effect, symptom severity was rated on a 5-point Likert scale ranging from,, 0: none at all” to,, 4: very strong”. An overall side effect score was obtained by summing scores for all items.

### Cold pressor test

The CPT ^18,68^ was performed using a refrigerated laboratory water bath (WCR-P22, Witeg, Germany, volume 22 l, temperature precision: ± 0.2 °C). Water was constantly circulated at a rate of 15 l/min to avoid the formation of a warm-water layer around the skin. To simulate a weak analgesic treatment effect ^18,19,69^, temperatures were covertly increased between the first and second CPT for all groups (including the non-treatment group); the pre-treatment CPT was performed at 6.0 ± 0.2 °C, the post-treatment CPT at 8.0 ± 0.2 °C.

Participants were asked to immerse the unclenched, non-dominant hand up to the wrist-crest, which was marked with a permanent marker. Participants could terminate testing anytime. They were asked to retract the hand from the water-bath and to provide the maximum VAS pain rating of 100 points if the pain became unbearable. Participants were informed that there was a time-limit for the test, but the exact maximum duration was not provided. Upon immersion, the experimenter logged the start of continuous VAS using a computerized trigger (button press). When the 180 second time-limit was reached, recordings were stopped automatically, and the experimenter asked the participant to withdraw the hand. When participants hand took their hand out of the bath earlier, the experimenter logged the end of recordings by a second button press.

Continuous ratings were obtained during CPT with a mechanical slider (11 cm, custom construction, sampling rate: 10 Hz), linked to an un-ticked 101-point VASs shown on screen (endpoints:,, 0: no pain”,,, 100: unbearable pain”). A laptop equipped with an external screen, *Matlab 2015b* (MathWorks, Natick, USA), and the *Psychtoolbox* (v 3) ^70^ was used to record participant’s ratings and to log experimental timings. Continuous heart rate (HR) recordings were obtained from the ring finger of the dominant hand using a standard bedside monitor (*Infinity Delta*, Dräger, Germany) equipped with a pulse spectrophotometer. The investigators logged starting and termination of CPT in HR-recordings by eliciting an separate electronic trigger signal via button press.

### Statistical analysis

Analysis was performed with *Matlab 2020b* and the *Statistics and Machine Learning* toolbox. Individual pain sensitivity in the CPT was the primary outcome and measured as percent-area-under-the-pain-curve (%AUPC) according to^16,30^. AUPC is an established summary metric for continuous CPT pain ratings and has been shown to be sensitive for detecting opioid analgesia^16,71,72^. Individual %AUPC vales were calculated based on the CPT pain rating curves (see: Fig. S1). Each %AUPC value corresponds to the average pain rating (AUPC_mean_) given over the full duration of a CPT, divided by the maximum possible AUPC value (AUPC_max_ = 100 VAS units * 180 seconds). %AUPC values quantitatively correspond to unstandardized AUPC values (unit: VAS-rating*ms) but have the advantage of being dimensionless and thus more intuitive to interpret. For participants who terminated testing early, the maximum VAS rating (100 units) was carried forward to 180 seconds^16^, so that participants who did not terminate testing and participants who terminate early can be analyzed as one sample, without requiring sub-partitioning of the sample. Still, CTP-tolerance time, i.e. the period that participants endured the CPT before retracting their hand from the water bath, was explored as a secondary outcome in participants terminating testing early, and AUPC_mean_ was explored as a secondary outcome in non-terminators.

Moreover, continuous pulse spectrophotometer-based HR recordings obtained before, during and after CPT were analyzed. The CPT is well known to induce a temporary increase in heart rate (HR) in healthy subjects^23,24^ and we aimed at estimating potential placebo treatment group effects on this CPT HR-response. For each participant, we calculated a pre-CPT-baseline as the mean HR 15 seconds preceding CPT. The secondary outcome measure CPT HR-response was calculated as the peak HR (HR-maximum) observed during the CPT period, minus the pre-CPT HR-baseline.

For all outcomes, the post-treatment timepoint was assessed as the dependent variable, which was tested for between-group factor of interest *group* (levels: no treatment, tasteless placebo, bitter placebo, sweet placebo) in a general linear model (GLM, as implemented in MATLAB’s *fitlm*). Where available, CPT pre-treatment baseline values were included amongst predictors as a co-variate, to account for inter-individual differences, as recommended for randomized trials with pre-treatment baselines^73^. Further, the fixed factor *Study* (levels: Sub-Study 1, Sub-Study 2, Sub-Study 3) was included to account for potential differences in sub-studies. We used robust parameter estimation (iteratively reweighted least-squares method, with *bisquare* weighting, the MATLAB default), instead of the ordinary-least-squares method to preclude potential outlier effects. GLMs were assessed in terms of variance explained (ANCOVA, *F*-test), followed-up by paired contrasts (*t*-tests on parameter estimates). In all F-and t-tests, the *p*-values for factor *group* (and its contrasts) were obtained via permutation testing (random Monte-Carlo permutation tests^74^, 5*10^5^ resamples) in order to relax the GLM assumptions of residual normality and homoscedasticity. One-sided p-values are reported for F-tests, two-sided p-values for t-tests. Unstandardized (b) and standardized (β) parameter estimates are provided with 95% confidence intervals (95% CI) obtained via bootstrapping (10^5^ resamples, BCa method, as implemented in MATLABs *bootci*).

The analyses performed were interpreted from a parameter estimation perspective, focusing on effect sizes, not p-values^75^. Standardized effect sizes were provided for all F-tests (partial eta^2^) and t-tests (β-estimates, see above). Treatment responder rates were calculated for the primary outcome %AUPC, in order to facilitate comparisons with clinical results: For this purpose, treatment responders were defined as the fraction of participants experiencing a pain reduction of >30%, relative to pre-treatment baseline, which is regarded as the level of practical significance in clinical pain^22^. Treatment responder rates (RRs) were thus calculated according as RR = ((AUPC_post_ – AUPC_pre_) / AUPC_pre_) < -.30), numbers needed to treat as the inverse of absolute risk reduction 1/(RR_GroupA_ -RR_GroupB_).

All main analyses were repeated in an intention-to-treat fashion, including all tested participants regardless of exclusion criteria, in order to allow for detecting deliberate selection bias. CPT-heart rate response, treatment expectation ratings, taste intensity ratings, and taste valence ratings were explored for associations with %AUPC to aid the interpretation of results. These covariates were centered and standardized for GLM analysis.

## Data Availability

The underlying data and analysis is available online at https://github.com/mzunhammer/analysis_placebo_taste.

## Supplementary Materials

### Supplementary Results

Fig. S1: continuous pain rating curves obtained in the cold pressor test

Fig. S2: time-effects over the course of sub-studies versus environmental temperatures

Fig. S3: single-subject heart-rate curves obtained in the cold pressor test

Fig. S4: peak heart-rate response during CPT versus % area under the pain curve

Fig. S5: time-course of taste intensity ratings in two small pilot studies

Fig. S6: pre- and post-CPT blood pressure, pooled across sub-studies

Table S1: sub-study sample features

Table S2: descriptive cold pressor test (CPT) results

Table S3: GLM results of % area under the pain curve, total sample

Table S4: GLM results of pain tolerance time, “terminator” sub-sample

Table S5: GLM results of pain tolerance time, “maxtimer” sub-sample

Table S6: Descriptive results heart rate, pooled across sub-studies

Table S7: GLM results of CPT HR-response, total sample

Table S8: descriptive results on treatment-related beliefs

Table S9: GLM results for % area under the pain curve versus taste-related ratings

Table S10: descriptive results blood pressure, pooled across sub-studies

## Acknowledgements

We thank Silke Bourdin, Katrin Forkmann, Christoph Ritter, Katrin Scharmach, and Frederik Schlitt for their assistance during data acquisition.

## Author contributions

Matthias Zunhammer: conceptualization, data analysis, writing the original draft; Gerrit Goltz and Maximilian Schweifel: data collection, review; Boris Stuck: conceptualization, methods, review; Ulrike Bingel: conceptualization, manuscript writing, review, supervision.

## Footnotes

1 For all participants, the only explicit mention of taste was made, when obtaining taste intensity and valence ratings *after* all other testing was completed

