## Supplement for "Savor the flavor — a randomized double-blind study on taste-enhanced placebo analgesia in healthy volunteers"

### Table of content

**Table of content 2**

**Supplementary Results 3**

**Supplementary Figures 5**

**Supplementary Tables 12**

**References 21**

### Supplementary Results

**Auxiliary analyses for the primary outcome: AUPC**

We noticed that there was a trend towards higher mean AUPC in Sub-Study 3. However, this sub-study effect was fully compensated by correcting for individual baseline (pre-treatment) AUPC values (see: Table 1 in the main manuscript). Nevertheless, we followed-up on this potential side-result of interest. Data for Sub-Study 3 was mostly obtained in the summer. Therefore, we explored a potential seasonal effects and found that AUPC was positively associated with higher outside temperatures (Fig. S2), indicating that individuals are more sensitive to cold water in the summer.

**Secondary outcome: pain tolerance in participants terminating CPT early**

In the per-protocol sample, 158 out of 297 (53.2%) of participants voluntarily terminated pre- or post-treatment CPT before the 180-second maximum duration. CPT tolerance time was assessed as secondary outcome in these "early terminator" participants. The same basic GLM used for the primary outcome was applied for tolerance time (Table S4), but tolerance time was log-transformed beforehand, to satisfy the assumption of a normal residual distribution. Contrast analysis indicated that placebo treatment prolonged CPT-tolerance time by +24.4%, 95% CI [12.0, 37.4] ($\beta = 0.36$ , 95% CI [0.17, 0.55], $t(151) = 3.37$, $p = .0010$) compared to no treatment in this sub-sample. Of note, these increases in tolerance time indicate a reduced pain and thus a placebo (not nocebo) effect. The effects of flavored, compared to neutral placebo on pain tolerance time () showed similar directions of effect and effect sizes as the primary outcome %AUPC, but were less robust in statistical terms, which is likely due to the smaller size of the "early terminator" sub-sample. Again, sweet placebo tended to show a more pronounced effect on tolerance time than both bitter and neutral placebo, however the contrast estimates comparing flavored placebo groups were far from reliable (Table S4).

**Secondary outcome: average pain ratings in participants not terminating CPT**

Moreover, average pain ratings were assessed as secondary outcome in the sub-group of 139 participants who endured the CPT for the full 180-second maximum duration in both pre- and post-treatment CPT. Again, the same GLM used for the primary outcome was applied. In the "maximum duration" sub-sample, placebo treatment was found to decrease average pain rating by an estimated -5.4 points VAS, 95% CI [-10.1, -0.02] ($\beta = 0.29$, 95% CI [-0.53, -0.01], $t(132) = -2.29$, $p < .0001$) compared to no treatment. Placebo treatments with added taste reduced average pain rating by an estimated -3.4 points VAS, 95% CI [-8.13, 1.72] ($\beta = 0.18$, 95% CI [-0.43, 0.10], $t(132) = -1.35$, $p = .162$) compared to neutral-tasting placebos. Again, sweet placebo showed a marginally stronger effect compared than bitter placebo (Table S5).

Taken together, CPT tolerance time in the early terminator sub-sample and average pain ratings in the maximum duration sub-sample showed similar directions of effect and effect sizes as AUPC, the primary outcome. Due to the smaller sub-sample sizes, analyses for CPT tolerance time and average pain ratings were associated with more uncertainty compared to the full sample.

**Auxiliary analyses for the secondary outcome: CPT peak heart rate response**

Effect size estimates and results for CPT peak heart rate response were confirmed when repeating analysis with *all* participants tested, even those fulfilling the pre-defined exclusion criteria (Table S7), which largely excludes that deliberate selection bias affected this outcome. Further, we assessed potential placebo effects on baseline-HR: At the start of the experiment (pre- treatment, pre-CPT) the mean HR-baseline was 79.7 ± 12.7 bpm across all groups (Table S6) and a GLM-analysis (controlling for sub-study) indicated no prominent group differences ($Df_1 = 3$, $Df_2 = 274$, $F= 0.77$, $p = .514$, partial $eta^2 = 0.008$). The mean HR-baseline after treatment was 73.7 ± 11.4 bpm, indicating that HR generally slowed from pre- to post-treatment (-6.0 bpm, 95% CI[5.04, 7.03], $t(279) = 11.9$, $p < .0001$), which is expectable since participants were sedentary over a continuous time. Again, post-treatment HR-baseline (controlling for pre-treatment pre-CPT HR-baseline and sub-study) showed no apparent group differences ($Df_1 = 3$, $Df_2 = 273$, $F= 0.99$, $p = .397$, partial $eta^2 = 0.011$), which suggests that placebo treatment did not have a detectable effect on baseline-HR before CPT.

Of note, there was no appreciable relationship between peak CPT HR-response and %AUPC (see: Figure S4) suggesting that peak CPT HR-response are not a surrogate marker of pain, replicating previous findings [1,2]. Interestingly, an exploratory analysis of questionnaire data indicated that peak CPT HR-responses were positively correlated with the "vigor" (0.26 bpm/score, 95%CI [0.04, 0.49], $\beta = 0.14$, 95% CI [0.02, 0.26], $t(276) = 2.32$, $p = .021$) and negatively correlated with the "depression" sub-scale (-0.29 bpm/score, 95%CI [-0.56, -0.02], $\beta = -0.13$, 95% CI [-0.25, -0.01], $t(276) = -2.09$, $p = .037$) of the profile of mood states questionnaire (POMS). These findings indicate that a stronger HR-response is weakly related to subjective, momentary, feelings of being "energized" and in a positive mood.

Finally, we assessed systolic and diastolic blood pressure measured before and after CPT (not measured continuously). However, no clear treatment group effects were found for these blood pressure measurements (see: Supplementary Results, Figure S6).

**Secondary outcome: blood pressure measurements**

The CPT typically induces an temporary increase spike in blood pressure (BP)[3,4]. In the present study we assessed treatment group effects on systolic and diastolic BP after CPT as a secondary outcome measure. In contrast to pain ratings and HR-recordings, BP was not measured continuously *during* CPT, but before and after CPT.

We could obtain valid BP-recordings from 292 out of the 297 participants of the per-protocol sample; 5 participants could not be analyzed due to missing recordings. Mean systolic and diastolic BP-curves during CPT are provided in Figure S6.

Pre-treatment, the mean pre-CPT BR-baseline was 125.4 ± 13.6 (systolic) / 74.3 ± 8.69 (diastolic) mmHg across all groups (Figure S6). A GLM-analysis (controlling for sub-study) suggested that baseline-BP showed no prominent differences between groups (systolic: $Df_1 = 3$, $Df_2 = 291$, $F= 1.03$, $p = .381$, partial $eta^2 = 0.011$, diastolic: $Df_1 = 3$, $Df_2 = 291$, $F= 1.30$, $p = .275$, partial $eta^2 = 0.013$). After treatment, the mean pre-CPT BP-baseline was 121.1 ± 12.3 (systolic) / 72.9 ± 9.40 (diastolic) mmHg, which indicates that baseline-BP tended to decline from pre- to post-treatment, which is plausible given that participants were subject to continuous sedentary time.

GLM-analysis of post-treatment pre-CPT BP-baseline (controlling for pre-treatment pre-CPT BP-baseline and sub-study) indicated no prominent group differences in post-treatment BP-baseline (systolic: $Df_1 = 3$, $Df_2 = 290$, $F= 1.90$, $p = .130$, partial $eta^2 = 0.019$, diastolic: $Df_1 = 3$, $Df_2 = 290$, $F= 0.77$, $p = .513$, partial $eta^2 = 0.008$) suggesting that placebo treatment did not have a sizeable effect on baseline-BP before the CPT-challenge.

A GLM testing effects of factor *group* on post-treatment post-CPT BP, controlling for post-treatment pre-CPT HR-responses, and fixed factor *study* indicated no prominent group differences in post-treatment BP (systolic: $Df_1 = 3$, $Df_2 = 286$, $F= 0.58$, $p = .628$, partial $eta^2 = 0.012$, diastolic: $Df_1 = 3$, $Df_2 = 285$, $F= 1.11$, $p = .347$, partial $eta^2 = 0.008$) suggesting that we could not find clear effects of placebo treatment on CPT-induced BP-changes.

This result was replicated in a GLM testing effects of factor *group* on post-treatment pre-to-post CPT changes in BP, *controlling for pre-treatment pre-to-post CPT changes in BP*, and fixed factor *study*. ANCOVA indicated no clear group differences in BP-changes over CPT after treatment, even if controlling for CPT-induced changes pre-treatment (systolic: $Df_1 = 3$, $Df_2 = 285$, $F= 0.94$, $p = .424$, partial $eta^2 = 0.010$, diastolic: $Df_1 = 3$, $Df_2 = 285$, $F= 0.59$, $p = .620$, partial $eta^2 = 0.006$).

**Supplementary Figures**

**Figures S1: continuous pain rating curves obtained in the cold pressor test**

**A**

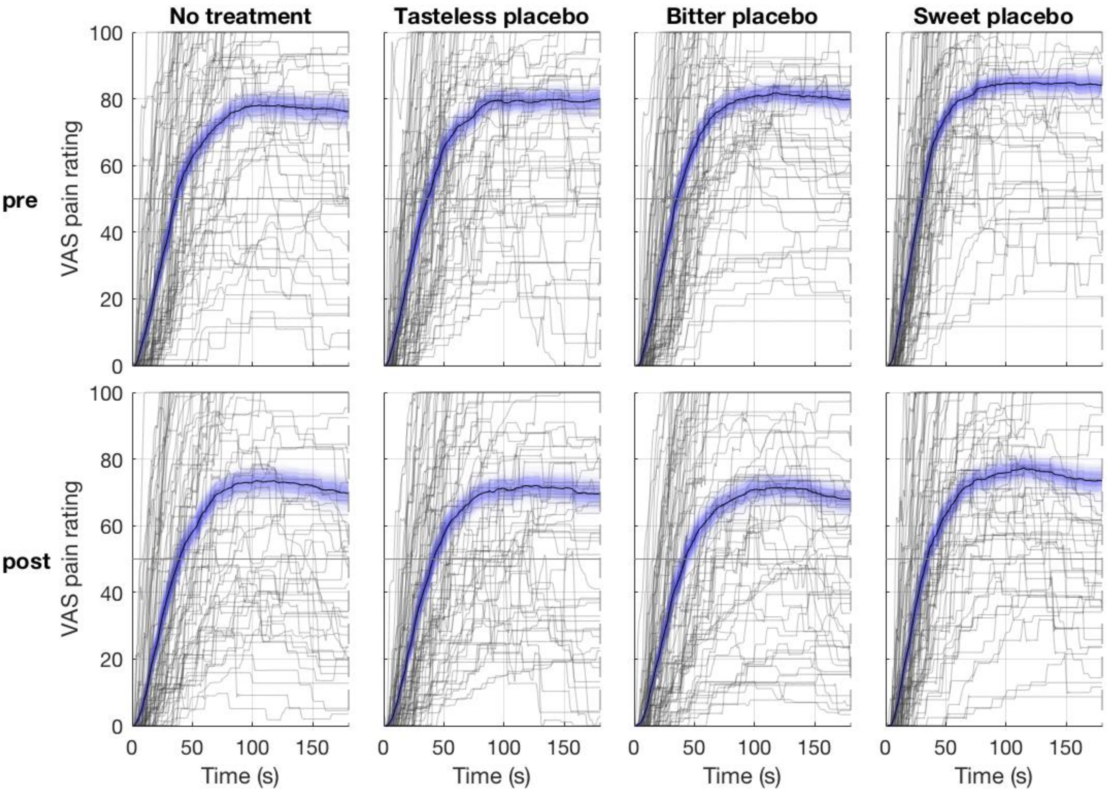


**B**

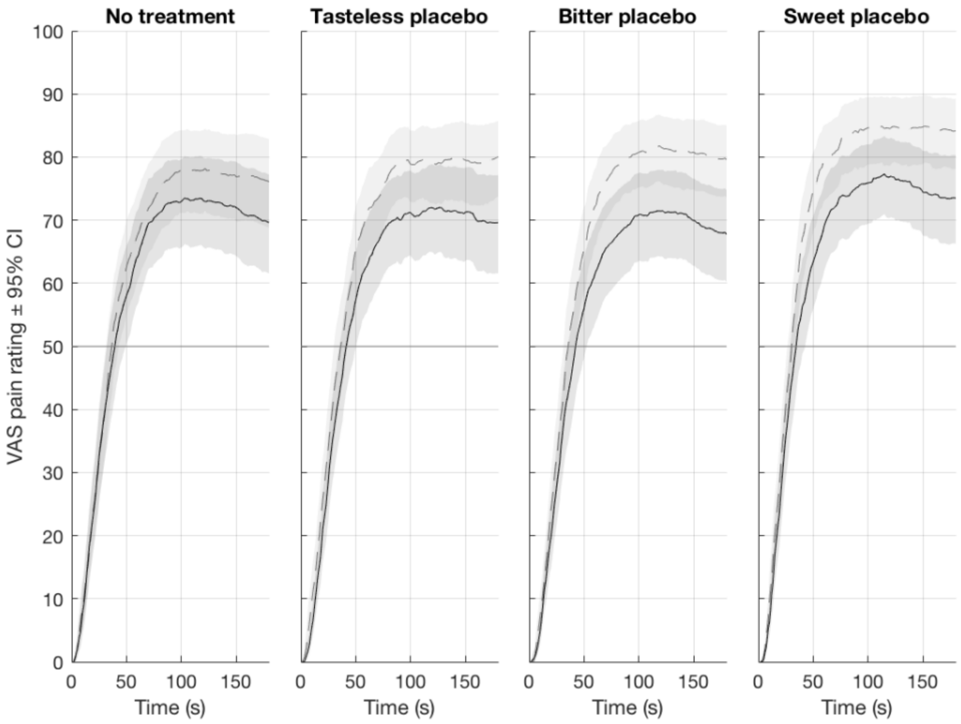


A) Grey lines denote single-subject (n = 297) pain rating curves. Bold black lines denote the mean rating curve for each condition, with blue shaded areas represents the corresponding bootstrapped 95% confidence interval. Curves of participants that ended testing before the 180-second time limit were carried forward with a VAS pain rating of 100.
B) Mean pain rating curves with bootstrapped 95% confidence intervals. Pre-treatment conditions are shown as dashed line + light grey area, post-treatment conditions as bold line + moderately grey area. Overlaps in 95% confidence interval are shown in dark grey.

**Figure S2: time-effects over the course of sub-studies versus environmental temperatures**

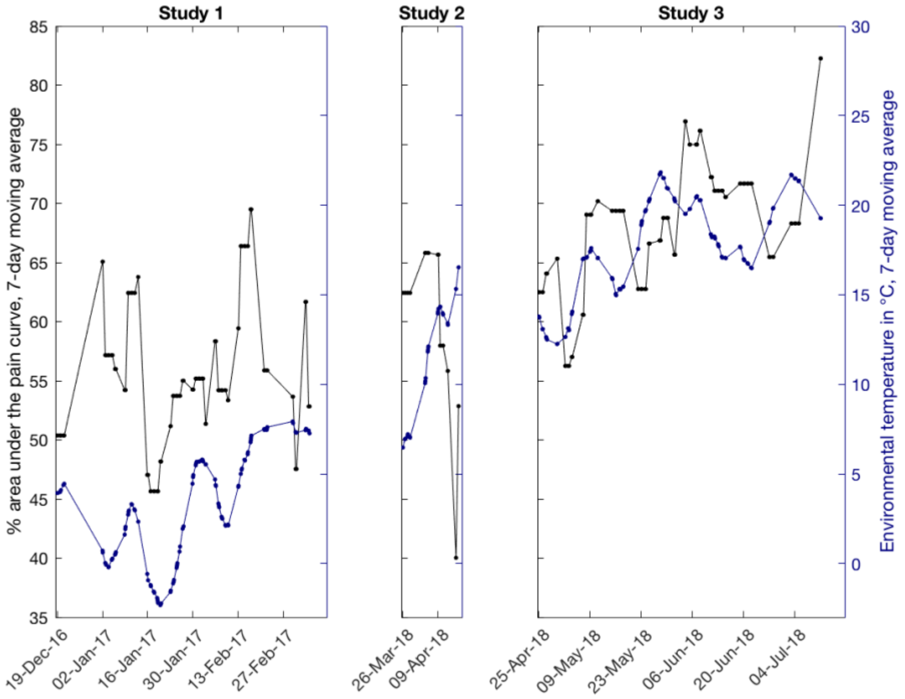


Participant's (n = 297) average (baseline and post-treatment) %AUPC values are shown on the left y-axes, respectively. Outside temperatures according to the German Weather Service (Düsseldorf station, hourly air temperature measures) are shown on the right y-axes. Note that the experiment involved only a brief acclimatization in the laboratory before baseline CPT-testing (22 minutes on average). Trends were visually enhanced by applying a 7-day moving average to all variables. CPT-tolerance times were inversed, for display purposes. Outside temperatures (unsmoothened) correlated significantly with both average %AUPC (r = 0.25, p<.0001), and average CPT tolerance time and (r = 0.25, p<.0001).

**Figures S3: single-subject heart-rate curves obtained in the cold pressor test**

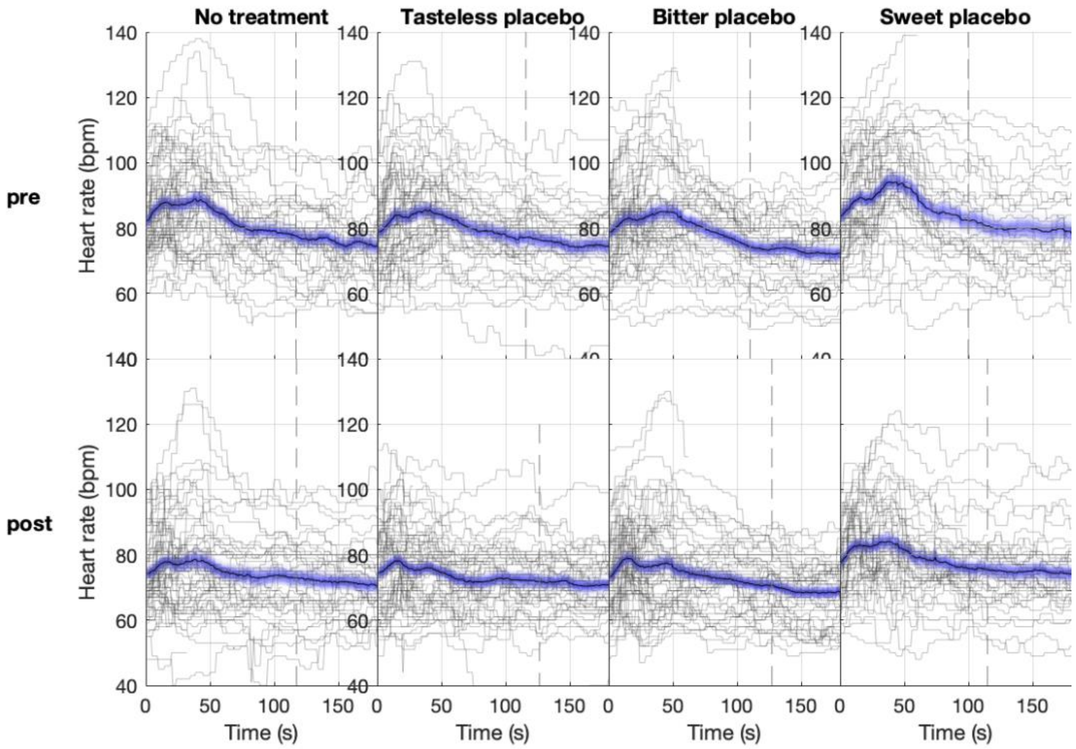


Grey lines denote all single-subject (n = 280) heart-rate curves from the per-protocol sample. Bold black lines denote the mean rating curve for each condition, with blue shaded areas represents the corresponding bootstrapped 95% confidence interval. Time = 0 denotes the start of CPT.

### Figures S4: peak heart-rate response during CPT versus % area under the pain curve

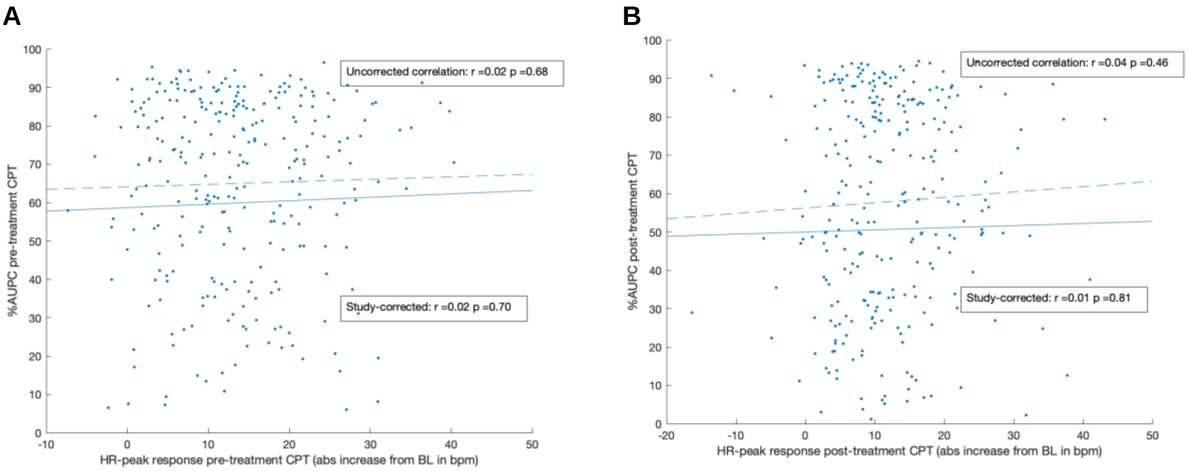


n = 280 (per-protocol sample with complete heart rate data). Solid lines denote simple linear regression of HR-peak response during the cold pressor test (CPT) versus %AUPC. Dashed lines denote the same relationship after correcting for sub-study averages. For the uncorrected relationship, Pearson's r is shown, for the sub-study corrected relationship it's the corresponding "rho" Value obtained from partial correlation analysis.

**Figure S5: time-course of taste intensity ratings in two small pilot studies**
**A**

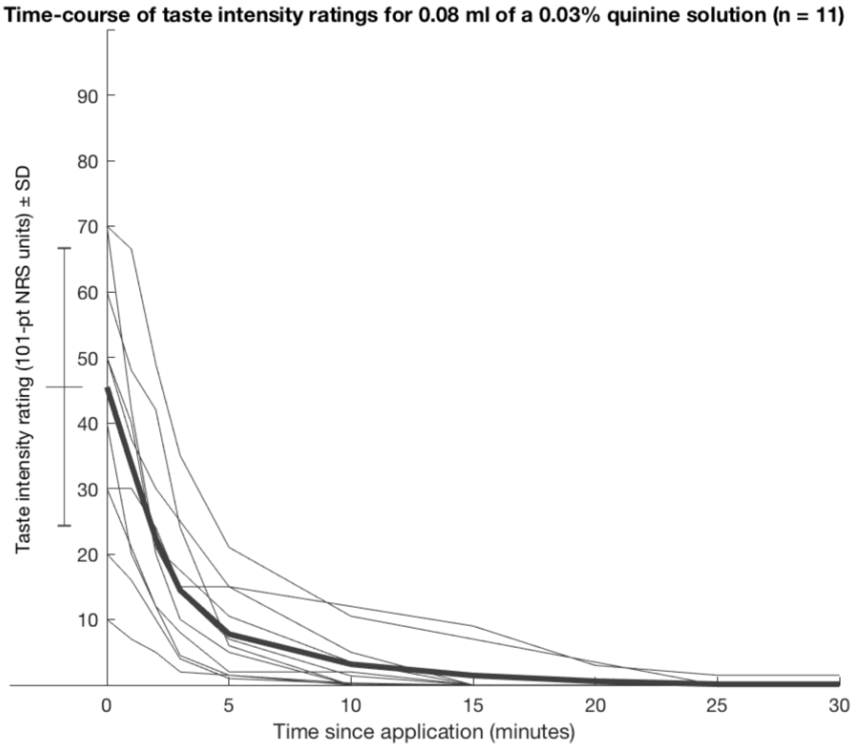


**B**

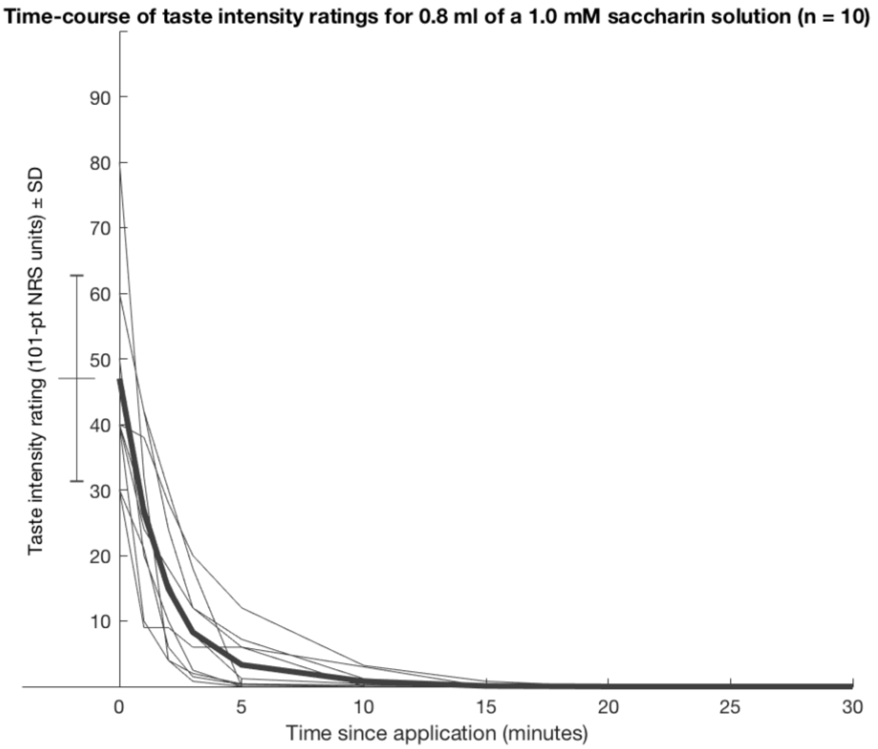


(A) Results of a pilot experiment (n = 11) testing the effects of 0.8 ml of a 0.03% quinine solution on taste intensity perception. (B) Results of a pilot experiment (n = 10) testing the effects of 0.8 ml of a 1.0 mM/l Saccharin solution on taste intensity perception. In both pilot experiments, taste intensity ratings were surveyed using a numeric rating scale (NRS) with endpoints 0: “no taste” and 100:”very intense”. The bar to the left of the y-axis depicts mean ± SD of taste intensity ratings at t = 0.

**Figure S6: pre- and post-CPT blood pressure, pooled across sub-studies**

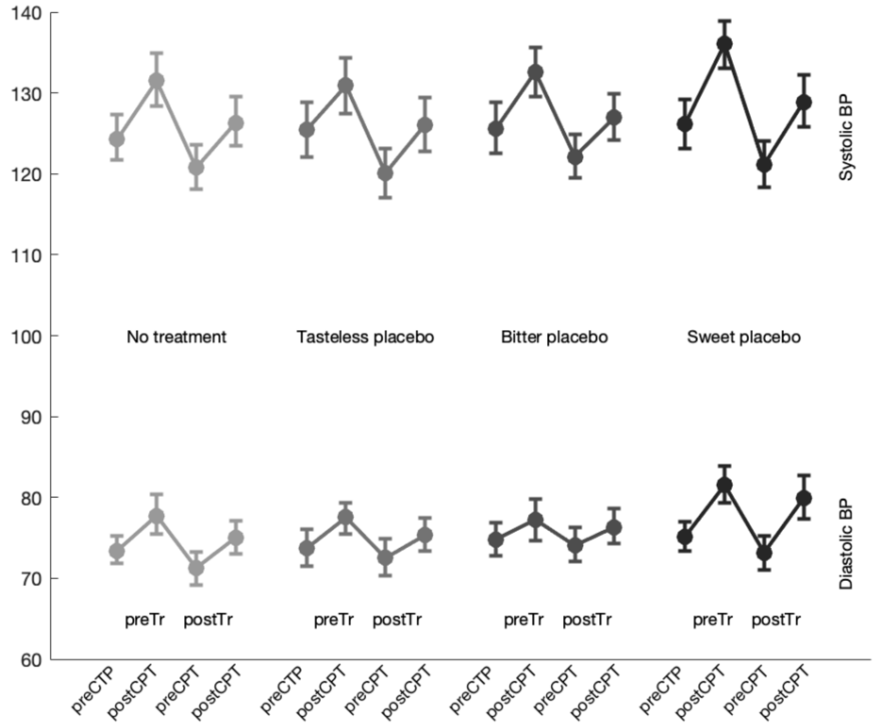


n = 292 (per-protocol sample with complete blood pressure recordings). Blood pressure measurements were taken before and after each cold pressor task.
Abbreviations: preTr: pre-Treatment, postTr: post-treatment

### Supplementary Tables

| | No treatment | Tasteless placebo | Bitter placebo | Sweet placebo | Total: |
|---|---|---|---|---|---|
| **Randomized (n)** | 46 | 46 | 46 | - | 138 |
| **Excluded (n)[a]** | 8 | 4 | 0 | - | 12 |
| **Per-protocol sample (n)[b]** | 38 | 42 | 46 | - | 126 |
| **Heart Rate Recordings (n)[c]** | 35 | 39 | 42 | - | 116 |
| **Sex (%male)** | 50% | 50% | 50% | - | 50% |
| **Age, mean (SD)** | 24.4 ± 2.6 | 25.0 ± 3.7 | 24.9 ± 3.0 | - | 24.7 ± 3.1 |
| **Handedness (%right)** | 87% | 88% | 98% | - | 91% |

**Table S1a: sample features of Sub-Study 1**

[a] Participants were excluded due to alcohol consumption (3), analgesic intake (3), participation in similar studies (2), insensitivity to CPT, i.e. pre-treatment %AUPC < 5% (3), chronic endocrine condition (1).

[b] An additional intention-to-treat analysis was performed with all available data.

[c] Number of participants where both pre- and post-treatment heart rate recordings could be analyzed.

| | No treatment | Tasteless placebo | Bitter placebo | Sweet placebo | Total: |
|---|---|---|---|---|---|
| **Randomized (n)** | 10 | 10 | 10 | - | 30 |
| **Excluded (n)[a]** | 1 | 1 | 0 | - | 2 |
| **Per-protocol sample (n)[b]** | 9 | 9 | 10 | - | 28 |
| **Heart Rate Recordings (n)[c]** | 9 | 9 | 10 | - | 28 |
| **Sex (%male)** | 56% | 56% | 50% | - | 54% |
| **Age, mean (SD)** | 23.6 ± 1.2 | 23.4 ± 1.6 | 25.0 ± 4.7 | - | 24.7 ± 3.1 |
| **Handedness (%right)** | 89% | 78% | 100% | - | 89% |

**Table S1b: sample features of Sub-Study 2**

[a] Participants were excluded due to alcohol consumption (1) and participation in similar studies (1).

[b] An additional intention-to-treat analysis was performed with all available data.

[c] Number of participants where both pre- and post-treatment heart rate recordings could be analyzed.

| | **No treatment** | **Tasteless placebo** | **Bitter placebo** | **Sweet placebo** | **Total:** |
|---|---|---|---|---|---|
| **Randomized (n)** | 25 | 25 | 25 | 75 | 150 |
| **Excluded (n)[a]** | 1 | 1 | 2 | 3 | 7 |
| **Per-protocol sample (n)[b]** | 24 | 24 | 23 | 72 | 143 |
| **Heart Rate Recordings (n)[c]** | 24 | 23 | 22 | 67 | 136 |
| **Sex (%male)[d]** | 46% | 33% | 52% | 59% | 51% |
| **Age, mean (SD)** | 24.6 ± 3.3 | 23.9 ± 3.1 | 23.9 ± 3.4 | 24.4±3.2 | 24.3 ± 3.2 |
| **Handedness (%right)** | 100% | 100% | 96% | 96% | 97% |

**Table S1c: sample features of Sub-Study 3**

[a] Participants were excluded due to psychiatric disorder (1), technical failure (1), vertigo after pre-testing CPT (2), participation in similar studies (2), insensitivity to CPT, i.e. pre-treatment %AUPC < 5% (1).

[b] An additional intention-to-treat analysis was performed with all available data.

[c] Number of participants where both pre- and post-treatment heart rate recordings could be analyzed.

[d] Note that group randomization for study 3 was performed without stratifying by sex to simplify recruitment. This decision was made after an interim analysis of sub-study 1, which suggested marginal effects of sex.

| | Pre- treatment | | | Post- treatment | | | Post – Pre Difference | | |
|---|---|---|---|---|---|---|---|---|---|
| | %AUPC | CPT duration | % maximum CPT duration* | %AUPC | CPT duration | % maximum CPT duration* | %AUPC | CPT duration | % maximum CPT duration* |
| **No treatment** | 63.4 ± 25.2 | 120.3 ± 65.2 | 52.1 ± 50.3 | 59.6 ± 27.6 | 119.8 ± 67.0 | 53.5 ± 50.2 | -3.8 ± 12.7 | -0.5 ± 29.4 | 1.4 ± 26.7 |
| **Tasteless placebo** | 63.7 ± 24.5 | 120 ± 65.5 | 49.3 ± 50.3 | 56.5 ± 27.2 | 129.0 ± 64.0 | 58.7 ± 49.6 | -7.2 ± 11 | 9.0 ± 22.7 | 9.3 ± 29.3 |
| **Bitter placebo** | 65.4 ± 23 | 116.8 ± 66.5 | 49.4 ± 50.3 | 56.4 ± 28.4 | 128.0 ± 66.0 | 60.8 ± 49.1 | -9 ± 11.6 | 11.2 ± 32.0 | 11.4 ± 32.0 |
| **Sweet placebo** | 70.4 ± 21.2 | 104.3 ± 66.3 | 40.3 ± 49.4 | 62.0 ± 25.5 | 120.8 ± 66.6 | 52.8 ± 50.3 | -8.4 ± 11.5 | 16.5 ± 37.5 | 12.5 ± 37.3 |

**Table S2: descriptive cold pressor test (CPT) results**
Total sample (n = 297); means ± standard deviations if not denoted otherwise. * Percentage of participants in the group enduring the full CPT duration of 180 seconds. † Obtained after CPT.

| Model Term | $Df_1$ | F | partial $eta^2$ | p* |
|---|---|---|---|---|
| Pre-treatment %AUPC | 1 | 2005 | 0.866 | < .001 |
| Study | 2 | 1.22 | 0.008 | .297 |
| Group | 3 | 6.85 | 0.062 | .0002 |

**Table S3a: GLM results of % area under the pain curve, total sample**

Full sample, no exclusions. n = 318. *p-Values for factor group are based on random (Monte Carlo) permutation testing. $df_2$ = 311, Adjusted $R^2$ = .873.

| Model Term | B [95% CI]* | Beta [95% CI]* | t | p** |
|---|---|---|---|---|
| Neutral Placebo > No Treatment | -3.56 [-6.72, -0.82] | -0.13 [-0.24, -0.03] | -2.28 | .058 |
| Bitter Placebo > No Treatment | -6.32 [-9.59, -3.27] | -0.23 [-0.35, -0.11] | -4.05 | .0007 |
| Sweet Placebo > No Treatment | -6.57 [-9.97, -3.36] | -0.24 [-0.36, -0.12] | -3.55 | .0006 |
| Bitter Placebo > Neutral Placebo | -2.76 [-6.02, 0.29] | -0.10 [-0.21, 0.01] | -1.78 | .139 |
| Sweet Placebo > Neutral Placebo | -3.02 [-6.38, 0.21] | -0.11 [-0.23, 0.01] | -1.64 | .109 |
| Sweet Placebo > Bitter Placebo | -0.25 [-3.57, 3.24] | -0.01 [-0.12, 0.12] | -0.14 | .895 |

**Table S3b: GLM contrasts of factor *group*, % area under the pain curve, total sample**

Full sample, no exclusions. n = 318. * 95% Confidence Intervals based on bootstrapping (asymmetric, BCa method). **p-Values are based on random (Monte Carlo) permutation testing. B denotes unstandardized model coefficients (unit: %AUPC), beta values show standardized model coefficients (units: SD). $df_2$ = 311, Adjusted $R^2$ = .873.

| Model Term | $Df_1$ | F | partial $eta^2$ | p* |
|---|---|---|---|---|
| Pre-treatment Tolerance Time | 1 | 306.1 | 0.669 | < .0001 |
| Study | 2 | 0.008 | <0.001 | .992 |
| Group | 3 | 3.991 | 0.073 | .009 |

**Table S4a: GLM results of pain tolerance time, "terminator" sub-sample**
Participants aborting either pre- or post-treatment CPT from per-protocol sample. Tolerance times (pre- and post treatment) had to be log-transformed for GLM analysis in order to achieve an approximate normal residual distribution. n = 158. *p-Value for factor group is based on random (Monte Carlo) permutation testing. $df_2 = 151$, Adjusted $R^2 = .681$.

| Model Term | B [95% CI]* | Beta [95% CI]* | t | p** |
|---|---|---|---|---|
| Neutral Placebo > No Treatment | +24.8% [8.76, 43.0] | 0.33 [0.12, 0.53] | 2.46 | .025 |
| Bitter Placebo > No Treatment | +24.5% [7.21, 49.2] | 0.32 [0.11, 0.60] | 2.46 | .023 |
| Sweet Placebo > No Treatment | +34.7% [12.4, 64.0] | 0.44 [0.17, 0.73] | 3.17 | .002 |
| Bitter Placebo > Neutral Placebo | -0.20% [-13.4, 19.7] | 0.00 [-0.21, 0.27] | -0.02 | .983 |
| Sweet Placebo > Neutral Placebo | +7.92% [-9.13, 30.5] | 0.11 [-0.15, 0.40] | 0.79 | .410 |
| Sweet Placebo > Bitter Placebo | +8.15% [-12.0, 30.9] | 0.12 [-0.19, 0.40] | 0.81 | .404 |

**Table S4b: GLM contrasts of factor *group,* pain tolerance time, "terminator" sub-sample**
Participants aborting either pre- or post-treatment CPT from per-protocol sample. Tolerance times (pre- and post treatment) were log-transformed for GLM analysis in order to approximate a normal residual distribution. n = 158. * B denotes unstandardized, back-transformed model coefficients (unit: exp(log(Tolerance Timer))-1,, 95% Confidence Intervals were obtained by bootstrapping (asymmetric, BCa method). **p-Values are based on random (Monte Carlo) permutation testing., beta values show back-transformed standardized model coefficients (units: SD). $df_2 = 151$, Adjusted $R^2 = . 681$.

| Model Term | $Df_1$ | F | partial $eta^2$ | p* |
|---|---|---|---|---|
| Pre-treatment %AUPC | 1 | 238.4 | 0.644 | < .001 |
| Study | 2 | 1.81 | 0.027 | .168 |
| Group | 3 | 2.48 | 0.053 | .064 |

**Table S5a: GLM results of pain tolerance time, "maxtimer" sub-sample**
Per-protocol sample, participants that did not abort pre- and post-treatment CPT from per-protocol sample. n = 158. *p-Value for factor group is based on random (Monte Carlo) permutation testing. $df_2 = 132$, Adjusted $R^2 = .635$.

| Model Term | B [95% CI]* | Beta [95% CI]* | t | p** |
|---|---|---|---|---|
| Neutral Placebo > No Treatment | -3.78 [-9.22, -1.70] | -0.20 [-0.47, 0.10] | -1.36 | .178 |
| Bitter Placebo > No Treatment | -6.76 [-12.5, 0.07] | -0.36 [-0.66, -0.01] | -2.46 | .015 |
| Sweet Placebo > No Treatment | -7.83 [-14.6, -0.43] | -0.42 [-0.80, -0.03] | -2.07 | .040 |
| Bitter Placebo > Neutral Placebo | -2.98 [-8.13, 2.61] | -0.16 [-0.43, 0.13] | -1.10 | .273 |
| Sweet Placebo > Neutral Placebo | -4.05 [-10.9, 2.97] | -0.22 [-0.57, 0.16] | -1.11 | .267 |
| Sweet Placebo > Bitter Placebo | -1.06 [-7.85, 5.77] | -0.06 [-0.42, 0.29] | -0.29 | .771 |

**Table S5b: GLM contrasts of factor *group*, "maxtimer" sub-sample**
Per-protocol sample, participants that did not abort pre- and post-treatment CPT from per-protocol sample. n = 158. * 95% Confidence Intervals based on bootstrapping (asymmetric, BCa method). **p-Values are based on random (Monte Carlo) permutation testing. B denotes unstandardized model coefficients (unit: %AUPC), beta values show standardized model coefficients (units: SD). $df_2 = 132$, Adjusted $R^2 = .635$.

| Group | No treatment | Taste-neutral placebo | Bitter placebo | Sweet placebo | Total: |
|---|---|---|---|---|---|
| n[a] | 68 | 71 | 74 | 67 | 280 |
| Heart rate pre-CPT baseline, pre-treatment, (bpm) | 80.5 ± 12.0 | 77.3 ± 12.7 | 78.3 ± 11.9 | 83.1 ± 13.5 | 79.7 ± 12.7 |
| Heart rate pre-CPT baseline, post-treatment, (bpm) | 74.7 ± 11.8 | 72.7 ± 12.2 | 71.4 ± 10.2 | 76.2 ± 10.8 | 73.7 ± 11.4 |
| Peak heart rate increase CPT, pre-treatment, (∆bpm) | +14.0 ± 8.8 | +14.1 ± 9.0 | +11.8 ± 9.1 | +14.8 ± 8.7 | +13.6 ± 8.9 |
| Peak heart rate increase CPT, post-treatment, (∆bpm) | +9.3 ± 8.9 | +10.7 ± 7.8 | +13.7 ± 9.0 | +13.7 ± 7.9 | +11.9 ± 8.6 |

**Table S6: descriptive results heart rate, pooled across sub-studies**

Abbreviations: bpm, beats per minute

[a] Number of participants where both pre- and post-treatment heart rate recordings could be analyzed. Reasons for additional exclusion compared to the per-protocol sample were: recording failure (n = 16), and extreme values (n = 1).

| Model Term | $Df_1$ | F | partial $eta^2$ | p* |
|---|---|---|---|---|
| Pre-treatment %AUPC | 1 | 58.7 | 0.168 | < .0001 |
| Study | 2 | 0.89 | 0.006 | .414 |
| Group | 3 | 5.98 | 0.058 | .00030 |

**Table S7: GLM results of CPT HR-response, total sample**

Total sample (including per-protocol exclusions w valid HR recordings) n = 297. *p-Values for factor group are based on random (Monte Carlo) permutation testing. $df_2$ = 290, Adjusted $R^2$ = .194.

| Model Term | B [95% CI]* | Beta [95% CI]* | t | p** |
|---|---|---|---|---|
| Neutral Placebo > No Treatment | 0.8 [-1.4 ,3.02] | 0.1 [-0.16 ,0.37] | 0.69 | 0.525 |
| Bitter Placebo > No Treatment | 4.54 [2.21 ,6.9] | 0.54 [0.27 ,0.83] | 3.89 | 0.00022 |
| Sweet Placebo > No Treatment | 2.71 [-0.17 ,5.21] | 0.32 [-0.02 ,0.63] | 1.97 | 0.035 |
| Bitter Placebo > Neutral Placebo | 3.75 [1.53 ,6.05] | 0.45 [0.18 ,0.71] | 3.22 | 0.0024 |
| Sweet Placebo > Neutral Placebo | 1.91 [-0.95 ,4.65] | 0.23 [-0.1 ,0.56] | 1.38 | 0.136 |
| Sweet Placebo > Bitter Placebo | -1.84 [-4.88 ,1.22] | -0.22 [-0.59 ,0.14] | -1.33 | 0.148 |

**Table S7: GLM contrasts of factor *group*, for CPT HR-response, total sample**

Full sample (including per-protocol exclusions w valid HR recordings) n = 297. * 95% Confidence Intervals based on bootstrapping (asymmetric, BCa method). **p-Values are based on random (Monte Carlo) permutation testing. B denotes unstandardized model coefficients (unit: beats-per-minute), beta values show standardized model coefficients (units: SD). $df_2$ = 290, Adjusted $R^2$ = .194.

| Group | No Treatment | Tasteless Placebo | Bitter Placebo | Sweet Placebo: |
|---|---|---|---|---|
| Treatment expectations[a] | NA | 44.8 ± 20.9 | 42.4 ± 20.5 | 38.2 ± 18.8 |
| Perceived treatment efficacy[b] | NA | 36.3 ± 25.4 | 34.9 ± 25.4 | 37.9 ± 28.1 |
| Treatment taste intensity[c] | NA | 7.6 ± 10.3 | 41.0 ± 27.2 | 20.0 ± 21.0 |
| Treatment taste valence[d] | NA | -3.6 ± 9.2 | -12.0 ± 17.5 | 12.1 ± 20.8 |
| Side effects[e] | NA | 0.5 ± 1.0 | 0.6 ± 1.1 | 1.2 ± 1.4 |

**Table S8: descriptive results on treatment-related beliefs**

[a] Expectations of pain relief, scale: 0, "no pain relief" to 100, "very strong pain relief".
[b] Rating of overall treatment efficacy, scale: 0, "no effect" to 100, "very strong effect".
[c] Rating of treatment taste intensity, scale: 0, "no taste" to 100, "very intense taste".
[d] Post-hoc rating of treatment taste valence, scale: -50, "very unpleasant taste" to 50, "very pleasant taste".
[e] Side effects, scale: sum-score of 18 five-point Likert-scale items (maximum possible score: 72). means ± standard deviations if not denoted otherwise.

| Model Term | $Df_1$ | F | partial $eta^2$ | p |
|---|---|---|---|---|
| Pre-treatment %AUPC | 1 | 1280 | .853 | < .001 |
| Pre-testing expectations of pain relief | 1 | 0.23 | .0002 | .635 |
| Post-testing ratings of treatment taste intensity | 1 | 0.00 | <.0001 | .978 |
| Post-testing ratings of treatment taste valence | 1 | 0.11 | .0001 | .739 |

**Table S9a: GLM results for % area under the pain curve versus taste-related ratings, placebo-treated groups**

$Df_2 = 221$

| Model Term | B ± SE | Beta ± SE | t | p |
|---|---|---|---|---|
| Pre-testing expectations of pain relief | -0.01 ± 0.03 | 0.00 ± 0.03 | -0.48 | .635 |
| Post-testing ratings of treatment taste intensity | -0.00 ± 0.03 | 0.00 ± 0.03 | -0.03 | .978 |
| Post-testing ratings of treatment taste valence | -0.01 ± 0.04 | -0.03 ± 0.03 | -0.33 | .739 |

**Table S9b: GLM contrasts of % area under the pain curve versus taste-related ratings, placebo-treated groups**

Model coefficients B show unstandardized effects in %AUPC, beta values show standardized effects in units of SD.

$Df_2 = 221$

| Group | No treatment | Taste-neutral placebo | Bitter placebo | Sweet placebo | Total: |
|---|---|---|---|---|---|
| n | 71 | 75 | 79 | 72 | 297 |
| Systolic blood pressure pre-CPT, pre-treatment | 124.3 ± 12.0 | 125.8 ± 15.4 | 125.5 ± 14.0 | 126.1 ± 12.8 | 124.3 ± 12.0 |
| Systolic blood pressure post-CPT, pre-treatment | 131.5 ± 14.2 | 130.8 ± 15.2 | 132.6 ± 13.6 | 136.1 ± 12.7 | 131.5 ± 14.2 |
| Systolic blood pressure pre-CPT, post-treatment | 120.8 ± 11.8 | 120.1 ± 13.3 | 122.2 ± 12.1 | 121.1 ± 12.2 | 120.8 ± 11.8 |
| Systolic blood pressure post-CPT, post -treatment | 126.3 ± 13.0 | 125.8 ± 14.5 | 127.1 ± 12.7 | 129.0 ± 13.7 | 126.3 ± 13.0 |
| Diastolic blood pressure pre-CPT, pre-treatment | 73.4 ± 7.5 | 73.6 ± 10.0 | 75.0 ± 9.2 | 75.2 ± 7.7 | 73.4 ± 7.5 |
| Diastolic blood pressure pre-CPT, pre-treatment | 77.7 ± 10.5 | 77.5 ± 8.4 | 77.5 ± 11.7 | 81.6 ± 9.8 | 77.7 ± 10.5 |
| Diastolic blood pressure pre-CPT, pre-treatment | 71.3 ± 8.9 | 72.5 ± 9.9 | 74.2 ± 9.7 | 73.3 ± 9.0 | 71.3 ± 8.9 |
| Diastolic blood pressure pre-CPT, pre-treatment | 75.0 ± 8.6 | 75.0 ± 8.8 | 76.4 ± 9.5 | 80.0 ± 11.4 | 75.0 ± 8.6 |

**Table S10: descriptive results blood pressure, pooled across sub-studies**

Abbreviations: CPT cold pressor test
